# Cholesterol Metabolism—Impacts on SARS-CoV-2 Infection Prognosis

**DOI:** 10.1101/2020.04.16.20068528

**Authors:** Yumeng Peng, Luming Wan, Chen Fan, Pingping Zhang, Xiaolin Wang, Jin Sun, Yanhong Zhang, Qiulin Yan, Jing Gong, Huan Yang, Xiaopan Yang, Huilong Li, Yufei Wang, Yulong Zong, Feng Yin, Xiaoli Yang, Hui Zhong, Yuan Cao, Congwen Wei

## Abstract

In this study, we specifically addressed the connection between the SARS-CoV-2 virus with host cholesterol metabolism. Plasma lipid profile was measured in 861 COVID-19 patients classified as mild (n=215), moderate (n=364), severe (n=217) or critical (n=65) and 1108 age- and sex-matched healthy individuals. We showed that the levels of both TG and HDL-C were significantly lower in patients with severe disease than in patients with moderate or mild disease. After successful treatment, cholesterol metabolism was reestablished in patients with SARS-CoV-2 infection. The serum concentrations of TC and HDL-C can be used as indicators of disease severity and prognosis in COVID-19 patients.

## Introduction

Since the outbreak of COVID-19 caused by severe acute respiratory syndrome coronavirus 2 (SARS-CoV-2), more than 10 million SARS-CoV-2 cases have been reported in over 200 countries (1). Cholesterol homeostasis is vital for proper cellular functions and is intimately associated with viral infections (2-4). Certain viruses manipulate plasma lipoproteins or plasma membrane lipid for its cell entry(5, 6). Although lipid pathophysiology and metabolomic analysis of SARS-CoV-2 infection showed massive metabolic suppression, the number of patients were limited (7, 8). Here in this study, lipid profiles were collected from 861 COVID-19 patients and 1108 age- and sex-matched individuals. We show that SARS-CoV-2 infection was associated with clinically significant lower level of HDL cholesterol (HDL-C), which can be used as indicators of disease severity and poor prognosis.

## Materials

### Clinical data sources

This study was approved by the Ethics Committee of the TaiKang Tongji Hospital ([2020] TKTJLL-003). As we collected the data from electronic medical files without contacting with the patients directly or analyzing the serum, written informed consent was waived. The basic informations and serum biochemical test results were collected from 861 COVID-19 patients in TaiKang Tongji Hospital (Wuhan, Hubei province, People's Republic of China) from February 16 to March 20, 2020. The median age was 60 years (range 16-96 years). Serum lipid profiles from 1108 age- and sex-matched individuals from the Third Medical Center of the Chinese PLA General Hospital were used as reference values. The median age was 61 years (range 31-96 years). The clinical and biochemical characteristics of these individuals were given in Table 1. The age and sex distributions did not differ between these two groups. Clinical manifestations were used to classify the disease status into four categories: mild, moderate, severe and critical. Mild clinical disease was characterized by mild symptoms with no pulmonary inflammation visible upon imaging. Disease was classified as moderate in the overwhelming majority of patients, who showed symptoms of respiratory infection, such as fever, cough, and sputum production, with pulmonary inflammation visible upon imaging. Disease was classified as severe when symptoms of dyspnea appeared, as assessed by the following symptoms: shortness of breath, respiration rate (RR) ≥ 30 bpm, resting blood oxygen saturation ≤ 93%, partial pressure of arterial oxygen/fraction of inspired oxygen ratio (PaO2/FiO2) ≤ 300 mmHg, or pulmonary inflammation that progressed significantly (> 50%) within 24 to 48 h. Critical disease was classified as the presence of respiratory failure, shock, or organ failure that required intensive care. Infection was confirmed in all patients by viral detection using quantitative RT-PCR, which simultaneously ruled out infection by other respiratory viruses, such as influenza virus A, influenza virus B, Coxsackie virus, respiratory syncytial virus, parainfluenza virus and enterovirus. All cases were diagnosed and classified according to the New Coronavirus Pneumonia Diagnosis Program (6th edition) published by the National Health Commission of China. Among the patients, 215 cases were classified as mild, 364 as moderate, 217 as severe and 65 cases as critical (Table 2).

### Statistical methods

In this study, GraphPad 6.0 software was used for statistical calculations and data plotting. All statistical analyses were performed using SPSS version 20 (SPSS, Chicago, IL, USA). Differences between two independent samples were evaluated by a t-test or Mann-Whitney U test. Differences between multiple samples were analyzed by one-way ANOVA followed by Bonferroni's post hoc analysis. Analysis of variance or the Kruskal-Wallis rank sum test was used for comparisons among multiple groups. The chi-square test was performed to compare count data. Significance values were set at: ns (not significant), p > 0.05; *, P < 0.05; **, P < 0.01; ***, P < 0.001.

## Results

Liver damage infected with SARS-CoV-2 prompted us to investigate whether infection with this virus could lead to alterations in lipid metabolism. The serum lipid levels of each subject showed that total TG levels were similar between the reference population and the COVID-19 patients (*P* =0.062; Table 1 and Figure 1A), but the serum TC and HDL-C levels were significantly lower in COVID-19 patients than in the reference population (*P* <0.0001 and *P* <0.0001, respectively; Table 1 and Figure 1B-C). Thus, SARS-CoV-2 infection was associated with clinically significant lower levels of TC and HDL-C. Interestingly, COVID-19 patients had significantly higher serum LDL-C levels than age- and sex-matched adults from the reference population (P < 0.0001; Table 1 and Figure 1D).

**Figure 1.**
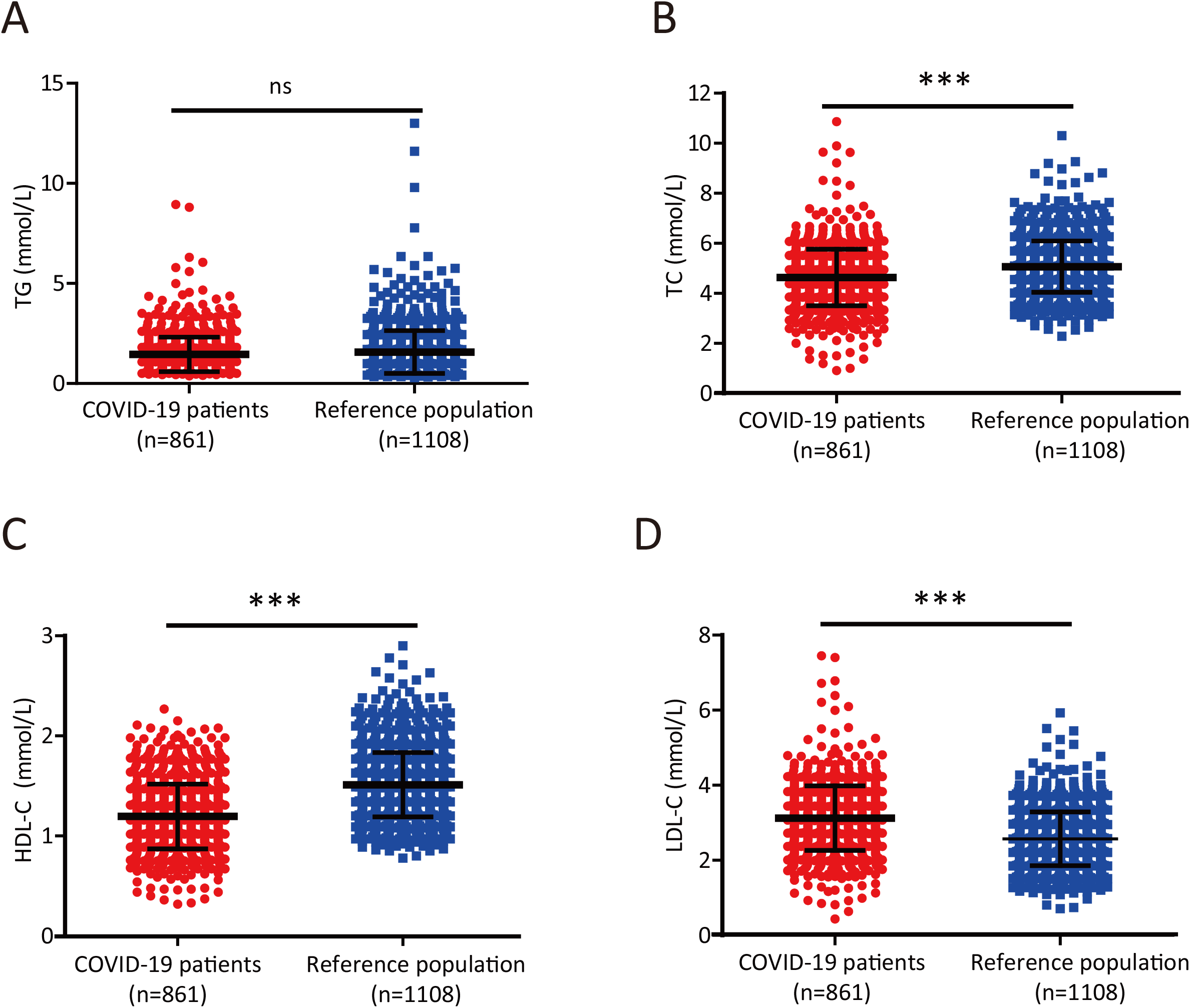
SARS-CoV-2 infection is associated with clinically significant lower levels of TC and HDL-C. (A-B) TG (A) and TC (B) levels in COVID-19 patients and reference subjects. Differences between the two groups were evaluated using the Mann-Whitney U test: ns, p > 0.05;*, P < 0.05; **, P < 0.01; ***, P < 0.001. (C-D) HDL-C (C) and LDL-C (D) levels in COVID-19 patients and reference subjects. Differences between the two groups were evaluated using the Mann-Whitney U test: ns, p > 0.05; *, P < 0.05; **, P < 0.01; ***, P < 0.001.

We next sought to determine whether serum lipid levels were correlated with disease conditions in COVID-19 patients. Although serum TG levels were significantly lower in patients with moderate disease than in those with mild disease (P< 0.0001; Table 2 and Figure 2A), they were similar in moderately, severely and critically ill patients (Table 2 and Figure 2A). However, the serum TC (Table 2 and Figure 2B), HDL-C (Table 2 and Figure 2C), and LDL-C (Table 2 and Figure 2D) concentrations were negatively correlated with disease severity and were particularly lower in critically ill patients than in patients with less severe disease. Specifically, the median HDL-C concentration in critically ill patients was 0.888 mmol/L (Table 2), which was below the normal range (1.03-1.89 mmol/L). Collectively, these results indicate that serum cholesterol levels may reflect the disease progression in patients infected with SARS-CoV-2.

We then selected 100 patients with severe symptoms but a cured outcome (survivors) and 16 with severe symptoms that died (non-survivors) and compared their lipid levels at admission (pretherapy) and after therapy (posttherapy). Although both groups of patients showed similar TG levels before and after therapy (Figure 3A), the TC and HDL-C levels of survivors rose significantly after therapy but dropped significantly in non-survivors (Figure 3B-C). LDL-C levels remained unchanged in survivors but decreased significantly after therapy (Figure 3D). In addition, we randomly selected 11 survivors and 9 non-survivors and monitored the dynamic changes in the HDL-C level of each patient from admission to recovery or death. Although the disease course differed in each patient, the HDL-C level increased progressively in 8 out of 11 survivors (Figure 3E), whereas the HDL-C level in all non-survivors showed a progressive decreasing trend (Figure 3F). Taken together, these results suggest that TC and HDL-C levels are prognostic predictors in patients with severe disease caused by SARS-CoV-2 infection.

**Figure 2.**
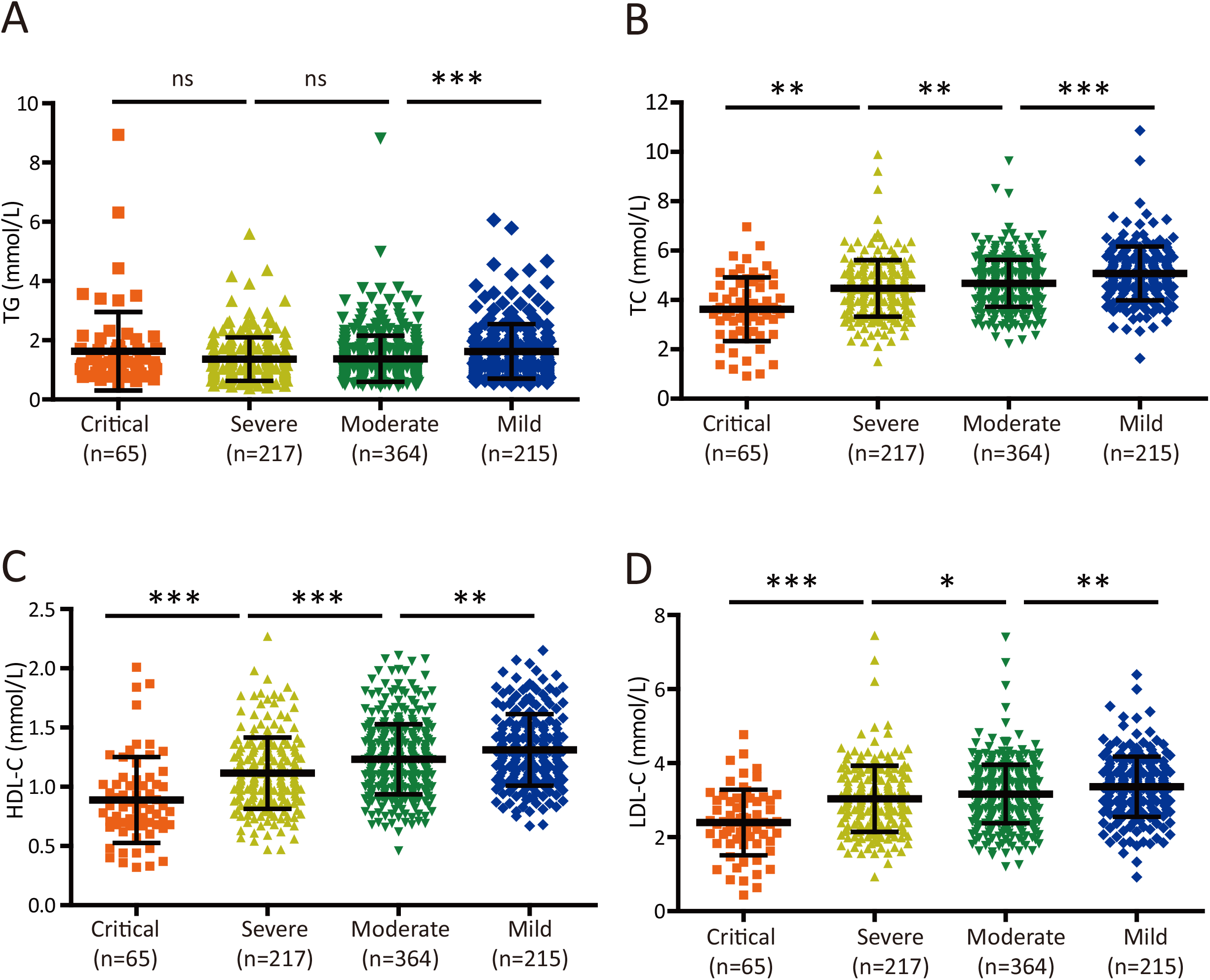
Cholesterol levels are inversely associated with disease severity in COVID-19 patients. (A-B) Plot of TG (A) and TC (B) levels in COVID-19 patients with four different levels of disease severity. Differences among groups were evaluated using the Mann-Whitney U test: ns, p > 0.05; *, P < 0.05; **, P < 0.01; ***, P < 0.001. (C-D) Plot of HDL-C (C) and LDL-C (D) levels in COVID-19 patients with four different levels of disease severity. Differences among groups were evaluated using the Mann-Whitney U test: ns, p > 0.05; *, P < 0.05; **, P < 0.01; ***, P < 0.001.

**Figure 3.**
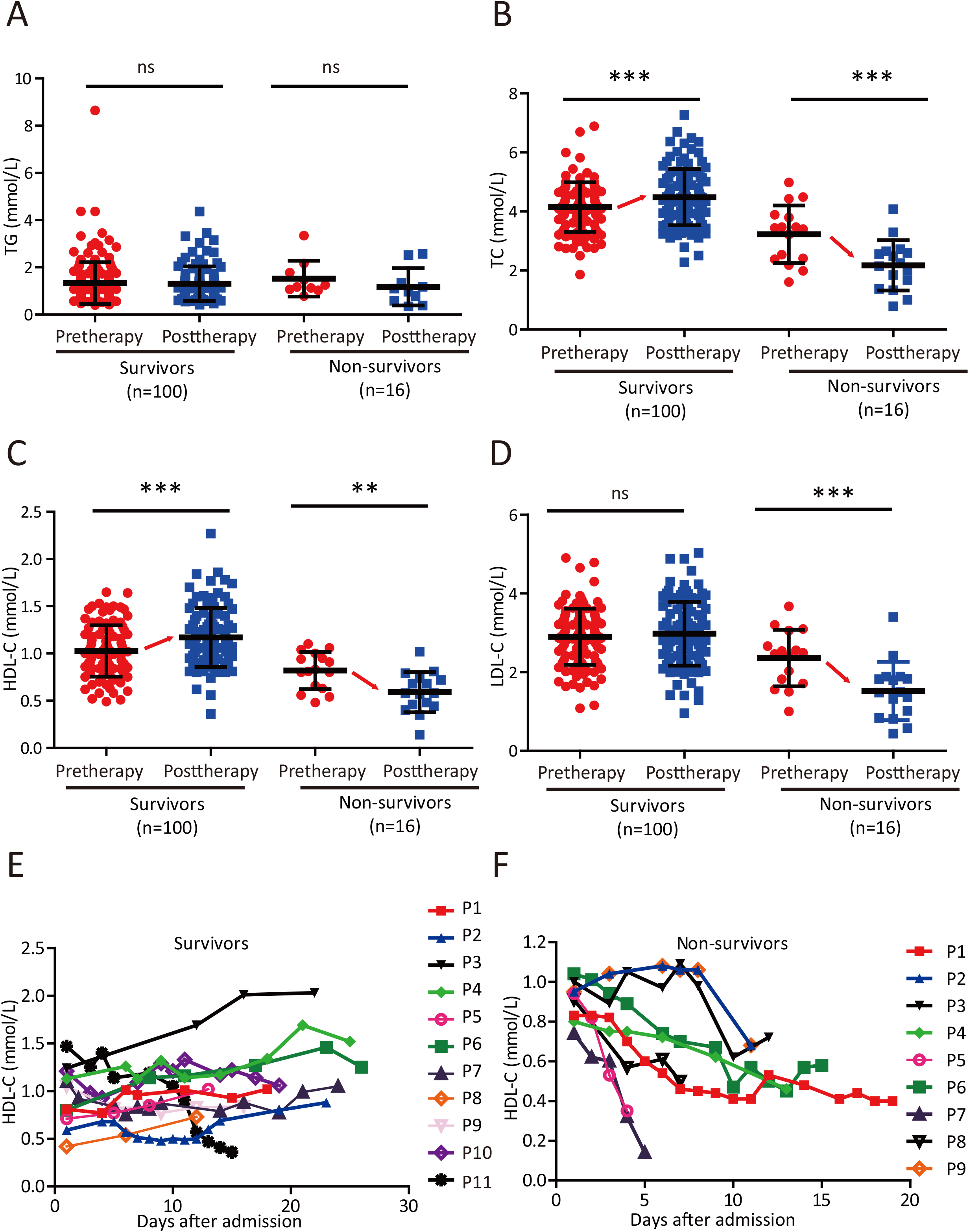
TC and HDL-C levels have prognostic value for COVID-19 patients with severe disease. (A-D) Changes in serum TG (A), TC (B), HDL-C (C) and LDL-C (D) levels in survivors and non-survivors before therapy (pretherapy) and after therapy (posttherapy). Differences between groups were evaluated using the Mann-Whitney U test: ns, p > 0.05; *, P < 0.05; **, P < 0.01; ***, P < 0.001.

## Discussion

Here in our study, although decreasing levels of LDL-C were associated with disease progression, COVID-19 patients had significantly higher levels of LDL-C than normal adults matched for age and sex. Our unpublished data confirmed this result in COVID-19 patients from another hospital in Hubei. More data are needed to reveal the mechanism underlying the association of increased LDL-C levels with COVID-19. Immune dysfunctions are a common feature in cases of SARS-CoV-2 infection and might be critical factors associated with disease severity (9). Here, we speculated that the immune-mediated cytokine storm might result at least partially from decreased HDL-C levels. HDL-C relieves cells of excessive amounts of cholesterol and has strong anti-inflammatory properties. In addition, HDL-C inhibits TLR-induced production of proinflammatory cytokines by macrophages (10). Moreover, Clinical studies haves shown that higher systemic levels of HDL and apoA-l are associated with less severe air flow obstruction (11). The correlation of reduced HDL-C levels with disease severity and mortality associated with SARS-CoV-2 infection implies the physiologically important function of HDL-C. HDL-C-mediated cholesterol efflux and selective cholesterol transport become dysfunctional in chronic metabolic diseases such as obesity or atherosclerosis, consistent with the increased mortality observed in elderly patients and patients with obesity or diabetes (12, 13). In addition, approximately half of the patients with SARS-CoV-2 infection had chronic underlying diseases, mainly cardiovascular and cerebrovascular diseases and diabetes (14). Therefore, intensive surveillance and early evaluation of the serum lipid levels in patients with preexisting conditions, especially in older patients with other comorbidities, might be needed.

## Data Availability

All data, models, or code generated or used during the study are available from the corresponding author by request.

## Author Contributions

All authors confirmed they have contributed to the intellectual content of this paper and have met the following 4 requirements: (a) significant contributions to the conception and design, acquisition of data, or analysis and interpretation of data; (b) drafting or revising the article for intellectual content; (c) final approval of the published article; and (d) agreement to be accountable for all aspects of the article thus ensuring that questions related to the accuracy or integrity of any part of the article are appropriately investigated and resolved.

Yuan Cao, financial support, provision of study material or patients.

## Conflict of interests

Nothing to disclose.

## Funding

This work is funded by the National Natural Science Foundation of China (No.81572620) and Shandong Provincial Natural Science Foundation of China (No.ZR2015HM003)

## Notes

### Competing Interest Statement

The authors have declared no competing interest.

### Clinical Trial

This is a single center based retrospective study, not a clinical trial.

### Summary of Updates

Article structure adjustment

